# Optimizing next-generation RSV prevention in Mali: a cost-effectiveness analysis of pediatric vaccination, maternal vaccination, and extended half-life monoclonal antibody immunoprophylaxis

**DOI:** 10.1101/2022.05.19.22275309

**Authors:** Rachel S. Laufer, Ranju Baral, Andrea G. Buchwald, James D. Campbell, Flanon Coulibaly, Fatoumata Diallo, Moussa Doumbia, Amanda J. Driscoll, Alison P. Galvani, Adama M. Keita, Kathleen M. Neuzil, Samba Sow, Clint Pecenka, Justin R. Ortiz, Meagan C. Fitzpatrick

**Affiliations:** Center for Vaccine Development & Global Health, University of Maryland School of Medicine, Baltimore, MD, USA; PATH, Seattle, WA, USA; Department of Pediatrics, Center for Vaccine Development & Global Health, University of Maryland School of Medicine, Baltimore, MD, USA; Centre pour le Développement des Vaccins, Ministère de la Santé, Bamako, Mali; Center for Infectious Disease Modeling and Analysis, Yale School of Public Health, New Haven, CT, USA

## Abstract

**Background:** Respiratory syncytial virus (RSV) is the most common cause of early childhood lower respiratory tract infection (LRTI) in low- and middle-income countries (LMICs). Maternal vaccines, birth-dose extended half-life monoclonal antibodies (mAbs), and pediatric vaccines are under development for prevention of respiratory syncytial virus (RSV) lower respiratory tract infection (LRTI) in young children.

**Methods:** We conducted an analysis of both health and economic impacts of RSV interventions used alone or in combinations in Mali. We modeled age-specific and season-specific risks of RSV LRTI in children through three years of life, using WHO Preferred Product Characteristics and data generated in Mali. Health outcomes included RSV LRTI cases, hospitalizations, deaths, and disability-adjusted life-years (DALYs). We identified the optimal combination of products across a range of scenarios.

**Finding:** We found that mAb delivered at birth could avert 878 DALYs per birth cohort at an incremental cost-effectiveness ratio (ICER) of $597 per DALY averted compared to no intervention if the product were available at $1 per dose. Combining mAb with pediatric vaccine administered at 10/14 weeks, 1947 DALYs would be prevented. The ICER of this combination strategy is $1514 per DALY averted compared to mAb alone. In an optimization analysis incorporating parameter uncertainty, mAb alone is likely to be optimal from the societal perspective at efficacy against RSV LRTI above 66%. The optimal strategy was sensitive to economic considerations, including product prices and willingness-to-pay for DALYs. For example, the combination of mAb and pediatric vaccine would be optimal from the government perspective at a willingness-to-pay above $775 per DALY. Maternal vaccine alone or in combination with other interventions was never the optimal strategy, even for high vaccine efficacy. The same was true for pediatric vaccine administered at 6/7 months.

**Interpretation:** At prices comparable to existing vaccine products, public health programs using extended half-life RSV mAbs alone or in combination with pediatric RSV vaccines would be impactful and efficient components of prevention strategies in LMICs such as Mali.

## INTRODUCTION

Respiratory syncytial virus (RSV) is the most common cause of early childhood lower respiratory tract infection (LRTI) in low- and middle-income countries (LMICs) [1]. There are no licensed RSV vaccines and only one licensed RSV monoclonal antibody (mAb), which is cost-prohibitive and programmatically unsuitable for most LMICs [2]. With new RSV prevention products in development [3], in 2016 the World Health Organization (WHO) Strategic Advisory Group of Experts on Immunization (SAGE) called for global policymaking for next-generation RSV vaccines and passive immunization [4]. Gavi, the Vaccine Alliance, announced plans in 2018 to support RSV immunization programs in eligible LMICs once products were licensed and recommended by the WHO [5].

Infrastructure for routine immunization in LMICs is most robust for children under 2 years of age [6]. In most Sub-Saharan African countries, routine immunization schedules include vaccinations at birth, 6 weeks, 10 weeks, 14 weeks, and 9 months [7]. Alternatives to pediatric RSV vaccine strategies include maternal RSV vaccination, which would use antenatal care delivery platforms in need of strengthening in many LMICs [8], or extended half-life mAbs, a technology new to LMIC immunization programs. The most practical strategy for RSV prevention in LMICs would be pediatric RSV vaccination programs integrated into established routine immunization schedules, but there are also challenges to this approach [9]. First, pediatric RSV vaccine research and development lags behind that for maternal vaccines and extended half-life mAbs [10]. Next, pediatric RSV vaccines are anticipated to require at least a two-dose series, with clinical trials assessing vaccination one month apart [11]. This increases logistic complexity and costs compared to the single-dose regimens expected for maternal vaccines and extended half-life mAbs. Finally, RSV vaccines may have lower efficacy during the first six months of life due to immune immaturity and interference from circulating maternal antibodies [9,12], a period coinciding with the highest RSV LRTI burden [13].

Studies assessing the health and economic impact of potential RSV prevention strategies are needed to inform immunization policy. While maternal vaccines and extended half-life mAbs may provide protection within the first six months of a child’s life, attack rates of RSV LRTI in low-income countries remain high through the first three years [1]. This suggests a role for longer-acting pediatric RSV vaccines and/or combinations of interventions to protect children through the longer period of risk. Among the potential products, the optimal RSV prevention strategy for LMICs is not clear. We adapted our previously validated health and economic impact model for RSV prevention in Mali to address this knowledge gap [14]. Mali has excellent RSV epidemiologic data collected over the last 10 years [1,15–17], it has high rates of RSV LRTI [16], and it is eligible for Gavi support [18]. We conducted an analysis of both health and economic impacts of RSV interventions used alone or in combinations through the first three years of life.

## METHODS

We evaluated the potential health and economic impacts of the following RSV LRTI prevention strategies in Mali, alone and in certain combinations: 1) one dose given at birth of an extended half-life mAb, administered seasonally; 2) one dose of maternal vaccine in the third trimester during routine antenatal visits; 3) two doses of pediatric vaccine given to infants during established 10- and 14-week (10/14 weeks) routine immunization visits; and 4) two doses of pediatric vaccine given to infants during established 6- and 7-month (6/7 months) routine immunization visits. We identified the optimal strategy within a net health benefits framework that incorporated empirical parameter uncertainty. All analyses were performed using R 4.0.2 (https://r-project.org). Model code is available at https://github.com/MCFitz/RSV-CEA-combos-Mali.git.

### Health and economic outcomes

We used a birth cohort model to simulate 12 monthly cohorts of infants from birth through the first 36 months of life [14]. We estimated RSV LRTI health outcomes under status quo (no RSV LRTI prevention) using Mali-specific epidemiological and economic data (Supplement Table 1, Supplement Figure 1). Probabilities of RSV LRTI depended on age-specific and season-specific incidence rates (Supplement Figure 2) [16]. Among infants under 6 months with RSV LRTI, we applied hospitalization rates observed in Mali [16]. To estimate hospitalization rates for older children, we applied a linear age-gradient as observed for children in the Gambia through 36 months, given the absence of reported hospitalization rates for children in Mali above 6 months of age. We used the RSV LRTI hospitalization rate in Mali for infants under 6 months as the starting value of the gradient (Supplement Figure 3) [13]. To estimate mortality, we sampled from age-specific case fatality rates (CFR) among infants hospitalized with RSV LRTI in LMICs as calculated previously [19], subset for ages 1 through 36 months (Supplement Figure 4). The CFR for infants from birth to six months of age in this study was similar to the attributable fatality rate calculated for Mali in the same age group (Supplement Figure 5).

We assigned a DALY value for each morbidity outcome using disability weights from the Institute for Health Metrics and Evaluation (Supplement Table 1, Supplement Figure 6) [20]. One DALY represents the loss of the equivalent of one year of full health. For each death, we calculated life-years lost using the life expectancy for Mali of 58 years and an annual discount rate of 3% [21].

For each intervention, we specified efficacy and duration of protection based on the WHO Preferred Product Characteristics (Table 1) [2,22]. Accordingly, the base-case efficacy against RSV LRTI was 70% for all products. Consistent with the Preferred Product Characteristics, we assumed that this efficacy pertains to pediatric RSV LRTI, and not upper respiratory illness or infection. The duration of protection was five months after administration for extended half-life mAb, four months after birth for maternal vaccine, and 12 months after second immunization with pediatric vaccine. We assumed no efficacy after only one dose of pediatric vaccine. We projected intervention coverage using 2019 Mali-specific data for routine immunization or antenatal care, respectively (Table 1). We assumed that pediatric and maternal vaccines would be delivered year-round, while mAb would be administered seasonally (Supplement Figure 7). The anticipated duration of pediatric vaccine protection supports a year-round immunization strategy. Uncertainties in gestational age for maternal vaccines targeting the third trimester of pregnancy, a high incidence of preterm birth, and low coverage of antenatal care are the rationale for a year-round maternal vaccination strategy. We used seasonal immunoprophylaxis of infants given stronger existing delivery platforms and fewer uncertainties than maternal vaccination.

**Table 1:** RSV LRTI prevention product characteristics used in primary analysis

|  | Administration | Efficacy against RSV LRTI<sup>1</sup> | Duration of protection<sup>1</sup> | Coverage<sup>2</sup> | Coverage rationale |
| --- | --- | --- | --- | --- | --- |
| Extended half-life monoclonal antibody | Single seasonal birth-dose | 70% | 5 months | 83.0% | Coverage for birth dose Bacillus Calmette-Guérin (BCG) vaccine in Mali |
| Maternal vaccine | Single dose given to pregnant women in third trimester | 70% | 4 months | 43.3% | The percentage of pregnant women attending at least four antenatal care visits (ANC4+) in Mali |
| Pediatric vaccine at 10 and 14 weeks | Two doses: first dose at 10 weeks and second dose at 14 weeks. Protection begins two weeks after the second dose | 70% | 12 months | 77.0% | Coverage for third dose diphtheria-tetanus-pertussis-containing vaccine (DTP3) in Mali |
| Pediatric vaccine at 6 and 7 months | Two doses: first dose at 6 months and second dose at 7 months. Protection begins two weeks after the second dose | 70% | 12 months | 70.0% | Coverage for first dose of measles containing vaccine (MCV1) in Mali |

1. From WHO Preferred Product Characteristics [2,22]
2. From WHO/UNICEF coverage estimates [31,38]
Note: cost-effectiveness ratios are not sensitive to intervention coverage.

We delineated economic costs, assessed in 2019 US dollars, by payer perspective. Medical treatment and hospitalization costs for RSV LRTI were based on costs of care for respiratory illness in Mali [17], and are the responsibility of the government [23]. We assumed the price of the intervention product, $1.00 per dose in the base case for all strategies reflecting the published cost for the pentavalent vaccine (DTP-HepB-Hib) [24], would be subsidized by a donor with the national government paying $0.20 per dose. The government would also cover associated procurement, storage, and administration costs, assumed to be $0.67 per dose, reflecting the mean incremental cost of adding one product to the established immunization schedule in low-income countries [25]. Although there are no current childhood immunization visits at 6/7 months of age in Mali, the planned rollout of the RTS,S malaria vaccine will add these time points [26].

For the base case scenario, we used the point estimate for each parameter to calculate the health and economic outcomes for all intervention strategies and status quo. To incorporate empirical uncertainty, we independently sampled 10,000 values from each parameter distribution (Supplement Table 1). We then simulated 10,000 expected health and economic outcomes for each intervention strategy and status quo, with confidence intervals representing 95% of simulated outcomes.

### Cost-effectiveness and Optimization

To evaluate cost-effectiveness, we first identified all strongly dominated strategies, defined as those for which another strategy provided greater health benefits at lower total cost. We then estimated the incremental cost-effectiveness ratio (ICER) as the ratio between the incremental DALYs averted by a strategy and the incremental cost incurred. For the least expensive non-dominated strategy, the incremental costs and benefits were calculated in comparison to status quo. For all other non-dominated strategies, the comparator was the next least-expensive non-dominated strategy. After calculating ICERs, we excluded any weakly dominated strategies, defined as those for which greater health benefit could be achieved at a lower ICER, and we recalculated ICERs for the remaining strategies accordingly. We conducted this analysis from the government and donor perspectives, including only the projected costs and benefits associated with each payer. We also evaluated the societal perspective inclusive of all projected direct costs and benefits.

We then conducted an optimization analysis to identify the optimal RSV LRTI prevention strategy across a variety of conditions. This optimization employed a net health benefits framework, where monetary costs were translated into their health value using willingness-to-pay for health (WTP) [27]. Net health benefits were calculated as:

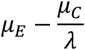

where *μ*_*E*_ represents the incremental health effect, *μ*_*C*_ represents the incremental cost, and *λ* represents the payer-specific WTP [28]. For our base case we specified WTP as $891 per DALY averted, the gross domestic product per capita in Mali. With each parameter set independently, we calculated the net health benefits associated with each intervention strategy. We identified the optimal strategy for a parameter set as that which provided the largest net health benefits. The probability that a strategy would be optimal was calculated as the number of parameter sets for which that strategy was optimal out of the total number of parameter sets.

### Sensitivity analyses

Given uncertainty regarding the appropriate WTP [29,30], we conducted a sensitivity analysis evaluating the optimal strategy choice across a range of WTP values ($0 to $7000 per DALY). In addition to the base case societal perspective, we performed this sensitivity analysis from the government and donor perspectives to characterize the potentially differential value of these interventions for each specific payer. In another sensitivity analysis, we evaluated the optimal strategy choice across product prices ranging from $0 to $4 per dose.

We conducted two-way sensitivity analyses evaluating the optimal strategy choice across simultaneous changes in 1) extended half-life mAb efficacy and its price, 2) extended half-life mAb efficacy and pediatric vaccine efficacy, 3) the price of pediatric vaccine administered at 10/14 weeks and the price of extended half-life mAb, and 4) the price of pediatric vaccine administered at 10/14 weeks and its efficacy. All parameter values not being investigated for a particular analysis were sampled from their empirical distributions as for the base case.

Finally, since RSV vaccination may have reduced performance if administered before six months of age compared to later in life [8], we used one-way sensitivity analyses to assess the optimal strategy choice across a wide range of efficacy values for pediatric vaccine administered at 10/14 weeks. In the first of these analyses, we identified the optimal strategy across vaccine efficacy changes regardless of whether other strategies were being deployed. In the second of these analyses, we only changed pediatric vaccine efficacy when vaccines were used in combination with either extended half-life mAb or maternal vaccine to specifically consider the possibility of interference among these products.

### Alternate scenario analysis

We considered an alternate scenario where maternal vaccine coverage was 89.1%, equivalent to the percentage of pregnant women in Mali attending at least one antenatal care visit [31], and with vaccine delivered seasonally instead of year-round. In this scenario, only our assumptions for the maternal vaccine strategy were changed, all other strategies remained the same as in the base case.

## RESULTS

We modelled an annual birth cohort of 778,680 children in Mali. We estimated the RSV LRTI cases, hospitalizations, deaths, and associated DALY loss for the cohort followed through 36 months of age under status quo and a suite of intervention strategies (Table 2). Over 77,000 cases, 6800 hospitalizations, and 136 deaths would be expected for this cohort in their first 36 months of life under status quo, corresponding to 3854 DALYs lost. We projected that extended half-life mAb, maternal vaccine, pediatric vaccine given at 10/14 weeks, and pediatric vaccine given at 6/7 months could prevent 1300, 470, 2322, and 1152 hospitalizations and 31, 12, 44, and 19 deaths, respectively. These strategies would avert 878, 347,1247, and 558 DALYs, respectively. Of combination prevention strategies, extended half-life mAb combined with pediatric vaccine given at 10/14 weeks would have the greatest health impact, averting 3325 hospitalizations, 69 deaths, and 1947 DALYs in the cohort.

**Table 2:** RSV LRTI preventive intervention cost-effectiveness analysis from the societal perspective

|  | RSV LRTI cases | RSV LRTI Hospitalizations | RSV LRTI Deaths | DALYs averted compared to status quo | Medical costs | Intervention costs | Total costs | Incremental cost-effectiveness ratio |
| --- | --- | --- | --- | --- | --- | --- | --- | --- |
| Status quo | 77685<br>(52215, 86505) | 6816<br>(3573, 8988) | 136<br>(59, 206) | --- | \$1249382<br>(742201, 1546908) | --- | \$1249382<br>(742201, 1546908) | --- |
| Maternal vaccine | 74378<br>(49916, 82841) | 6346<br>(3330, 8349) | 124<br>(54, 186) | 347<br>(140, 569) | \$1176679<br>(699904, 1455074) | \$563469 | \$1740148<br>(1263373, 2018543) | *Weakly dominated* |
| Extended half-life monoclonal antibody | 68535<br>(45967, 76320) | 5516<br>(2896, 7252) | 105<br>(46, 157) | 878<br>(369, 1393) | \$1048241<br>(624702, 1288465) | \$724993 | \$1773233<br>(1349695, 2013457) | \$597<br>(354, 1621) |
| Pediatric vaccine at 10 & 14 weeks | 50285<br>(33969, 56 438) | 4494<br>(2363, 5945) | 92<br>(40, 141) | 1247<br>(548, 1848) | \$817669<br>(486934, 1018490) | \$2004024 | \$2821693<br>(2490958, 3022514) | *Weakly dominated* |
| Pediatric vaccine at 6 & 7 months | 59748<br>(40417, 67172) | 5664<br>(2992, 7512) | 117<br>(50, 179) | 558<br>(245, 805) | \$1006701<br>(596633, 1261419) | \$1821840 | \$2828541<br>(2418472, 3083259) | *Strongly dominated* |
| Maternal vaccine plus pediatric vaccine at 10 & 14 weeks | 46978<br>(31760, 52694) | 4024<br>(2122, 5326) | 80<br>(35, 121) | 1594<br>(705, 2387) | \$744966<br>(445934, 924936) | \$2567493 | \$3312459<br>(3013427, 3492429) | *Weakly dominated* |
| Maternal vaccine plus pediatric vaccine at 6 & 7 months | 56440<br>(38155, 63435) | 5194<br>(2745, 6890) | 104<br>(45, 158) | 906<br>(400, 1343) | \$933998<br>(554562, 1167781) | \$2385309 | \$3319307<br>(2939871, 3553090) | *Strongly dominated* |
| Extended half-life monoclonal antibody plus pediatric vaccine at 6 & 7 months | 50597<br>(34191, 56823) | 4364<br>(2305, 5772) | 85<br>(37, 129) | 1436<br>(630, 2155) | \$805560<br>(480774, 1002645) | \$2546832 | \$3352392<br>(3027606, 3549477) | *Strongly dominated* |
| Extended half-life monoclonal antibody plus pediatric vaccine at 10 & 14 weeks | 43224<br>(29178, 48452) | 3491<br>(1840, 4610) | 67<br>(29, 101) | 1947<br>(855, 2936) | \$662438<br>(398664, 819499) | \$2729016 | \$3391454<br>(3127680, 3548516) | \$1514<br>(999, 3705) |

Note: All results displayed in the table have been rounded to the nearest whole number. The 95% credible intervals are given in parentheses where applicable.

We delineated costs by payer for all interventions and status quo (Table 2, Supplement Table 2). From the societal perspective and at a product cost of $1.00, extended half-life mAb, maternal vaccine, and pediatric vaccine given at 10/14 weeks would have incremental costs of $524,000, $491,000, and $1,580,000, respectively, compared to status quo. Cost variation among these interventions was primarily due to their different schedules and coverage; for instance, we assumed two doses of pediatric vaccine would be delivered year-round, while extended half-life mAb would be a single dose delivered seasonally (Supplement Figure 7). The highest-cost combination strategy would be extended half-life mAb at birth plus pediatric vaccine given at 10/14 weeks, with an incremental cost of $2,060,000 compared to mAb alone.

In a scenario where all products met the WHO Preferred Product Characteristics and were all available in Mali, the lowest-cost non-dominated strategy would be extended half-life mAb, with an ICER of $597 per DALY averted (Figure 1, Table 2). The only other non-dominated strategy was the combination of extended half-life mAb followed by pediatric vaccine at 10/14 weeks, with an ICER of $1514 per DALY averted compared to extended half-life mAb alone (Figure 1, Table 2). Although maternal vaccination was the lowest-cost option due to modest expected uptake, extended half-life mAb was projected to avert DALYs more efficiently, and therefore maternal vaccination was weakly dominated in a situation where both products are available. Pediatric vaccine administered at 6/7 months was strongly dominated because it averted fewer DALYs and was more costly than extended half-life mAb.

**Figure 1:**
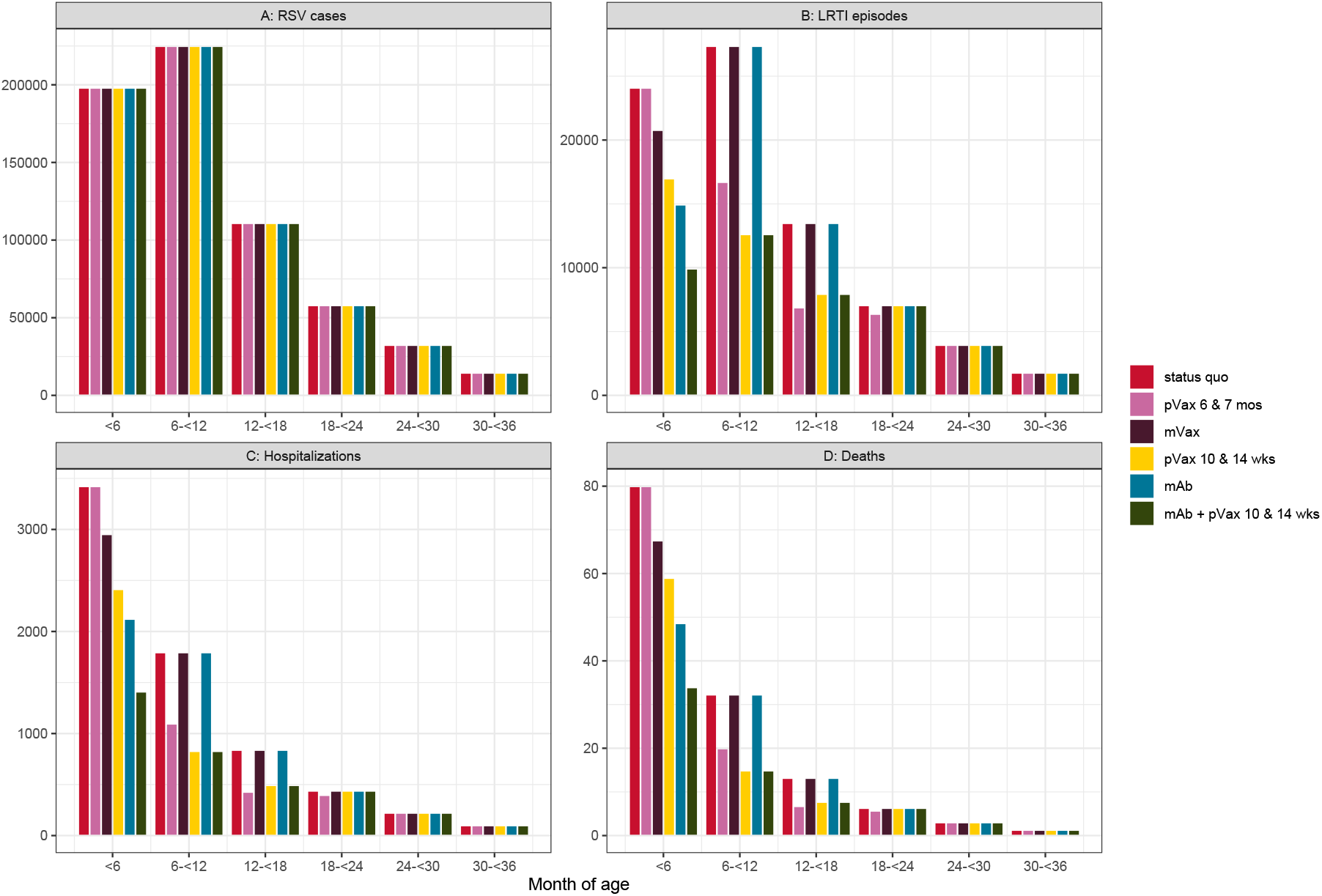
Health outcomes by age in months for a single year birth cohort in Mali, under status quo and modeled scenarios of RSV LRTI prevention: A) RSV cases, B) LRTI episodes, C) hospitalizations, and D) deaths.

We evaluated the probability that each prevention strategy would be optimal from the societal perspective across a range of product prices to identify potential price ceilings on RSV LRTI prevention products (Figure 3A). Above a product price of $1.24, no prevention strategy had a greater than 50% probability of being optimal. We then evaluated the probability that each prevention strategy was optimal from the societal perspective across a range of WTP, fixing product price at $1.00 based on the cost of pentavalent vaccine (Figure 3B). Below WTP of $750, status quo was most likely to be optimal. At WTP between $750-$1850, the preferred strategy was extended half-life mAb, and above WTP $1850 the preferred strategy became extended half-life mAb in combination with pediatric vaccine administered at 10/14 weeks. When the government perspective was considered, a similar progression was followed although with lower thresholds (Supplement Figure 8). Extended half-life mAb became optimal at WTP of $275 through $775, above which the optimal strategy was the combination of mAb and pediatric vaccine at 10/14 weeks. From the donor perspective, the progression was similar, with transition points at WTP of $500 and $1050 (Supplement Figure 8).

**Figure 2:**
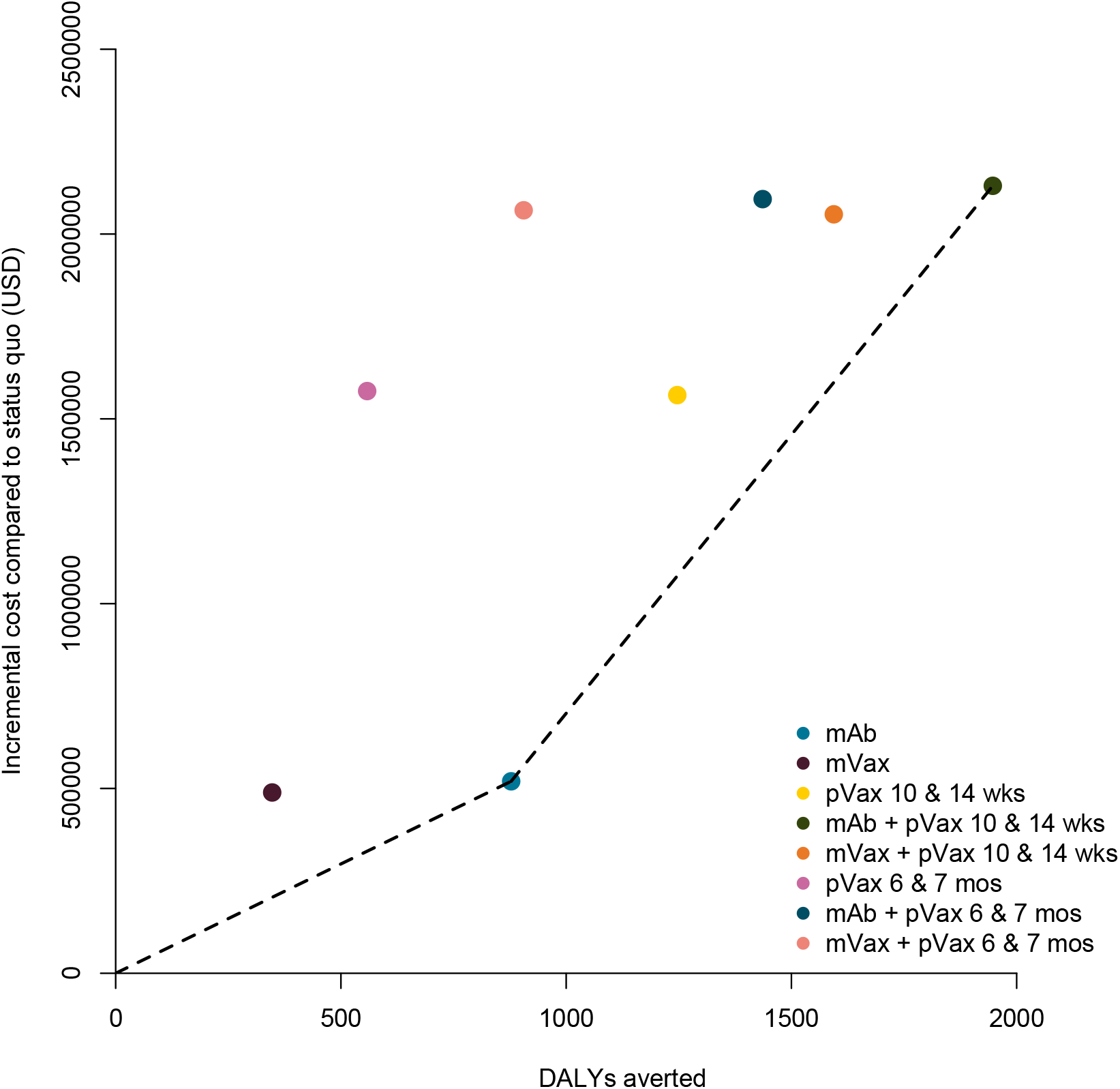
Additional cost per disability-adjusted life-year (DALY) averted for each RSV LRTI prevention scenario: extended half-life monoclonal antibody (mAb), maternal vaccine (mVax), pediatric vaccine (pVax) at both 10 & 14 weeks and 6 & 7 months, as well as combination strategies, mAb + pVax and mVax + pVax. The dashed line marks the efficient frontier, connecting all non-dominated interventions.

**Figure 3:**
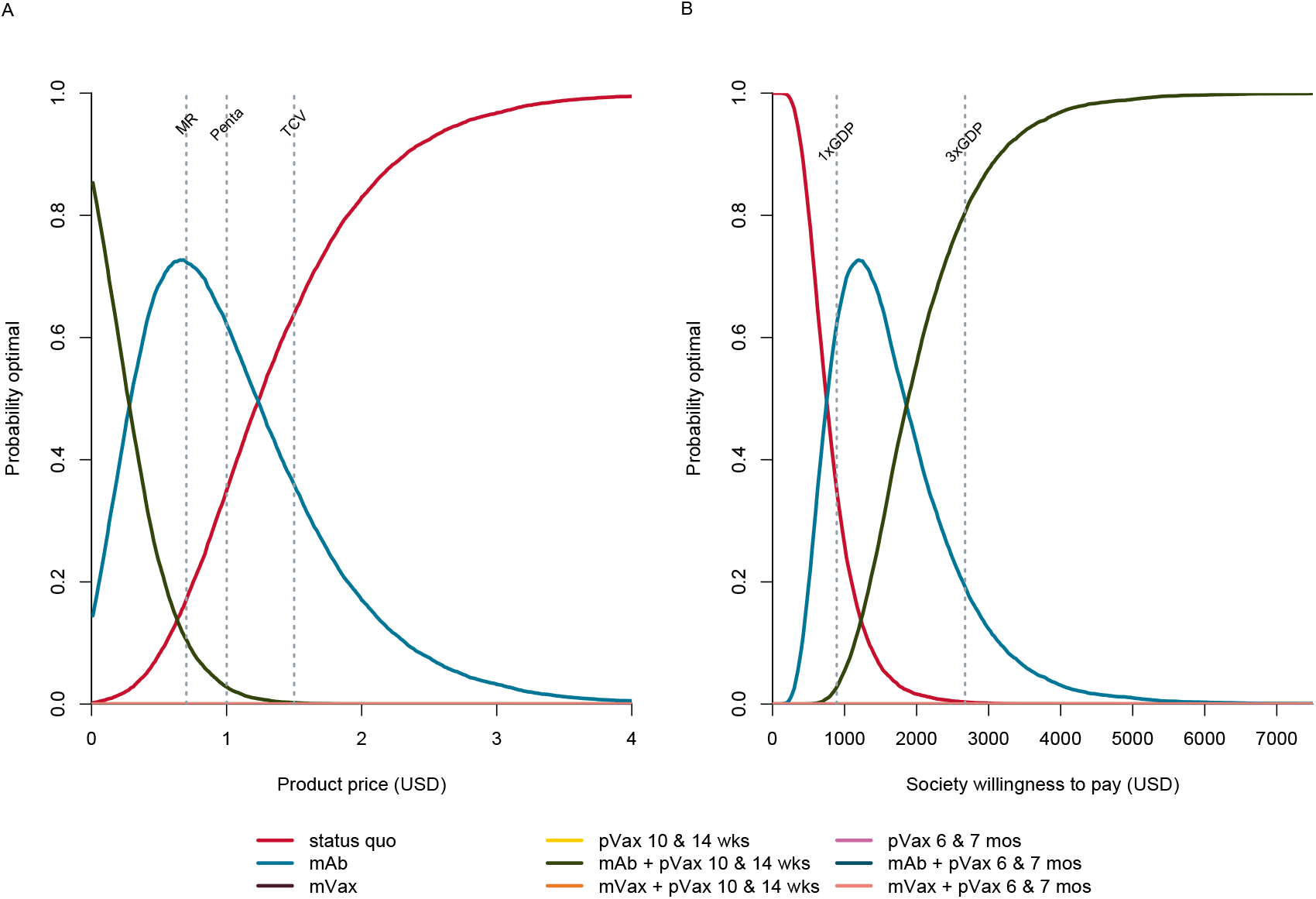
**A)** Probability each RSV LRTI prevention strategy is optimal given a product price per dose and willingness-to-pay of $891, equal to the per capita GDP of Mali. For comparison, prices for Measles-Rubella (MR), DPT Hep B Hib (Penta), and Typhoid (TCV) vaccine are indicated by the dashed lines [24,39]. **B)** Probability each RSV LRTI prevention strategy is optimal given a societal WTP to avert disability-adjusted life-years and an intervention product price of $1.00. Although they are no longer considered standard for WTP values, benchmarks of 1 X GDP and 3 X GDP per capita are drawn here for comparison. The competing scenarios are status quo, extended half-life monoclonal antibody (mAb), maternal vaccine (mVax), pediatric vaccine (pVax) administered at 10 and 14 weeks, pVax administered at 6 & 7 months, as well as combinations mAb + pVax and mVax + pVax. Strategies not visible on the plot are never the optimal choice and therefore remain at y = 0.

We evaluated the optimal strategy across changing pediatric vaccine efficacy when administered at 10/14 weeks (Supplement Figure 9). Even when pediatric vaccine has 100% efficacy, extended half-life mAb remains the strategy choice with the highest likelihood of being optimal.

We evaluated the optimal strategy across simultaneous changes in the efficacy and price of extended half-life mAb and pediatric vaccine at 10/14 weeks (Figure 4). As the efficacy of extended half-life mAb increased, mAb became more likely to be optimal, even at higher prices (Figure 4A). However, even as efficacy of pediatric vaccine approached 100%, it never became the optimal strategy (Figure 4B, 4D). Only with simultaneously high efficacy and low price did pediatric vaccine outcompete extended half-life mAb (Figure 4B). When pediatric vaccine cost less than $0.25 per dose and extended half-life mAb cost less than $0.46 per dose, sequential use of these two products became optimal (Figure 4C).

**Figure 4:**
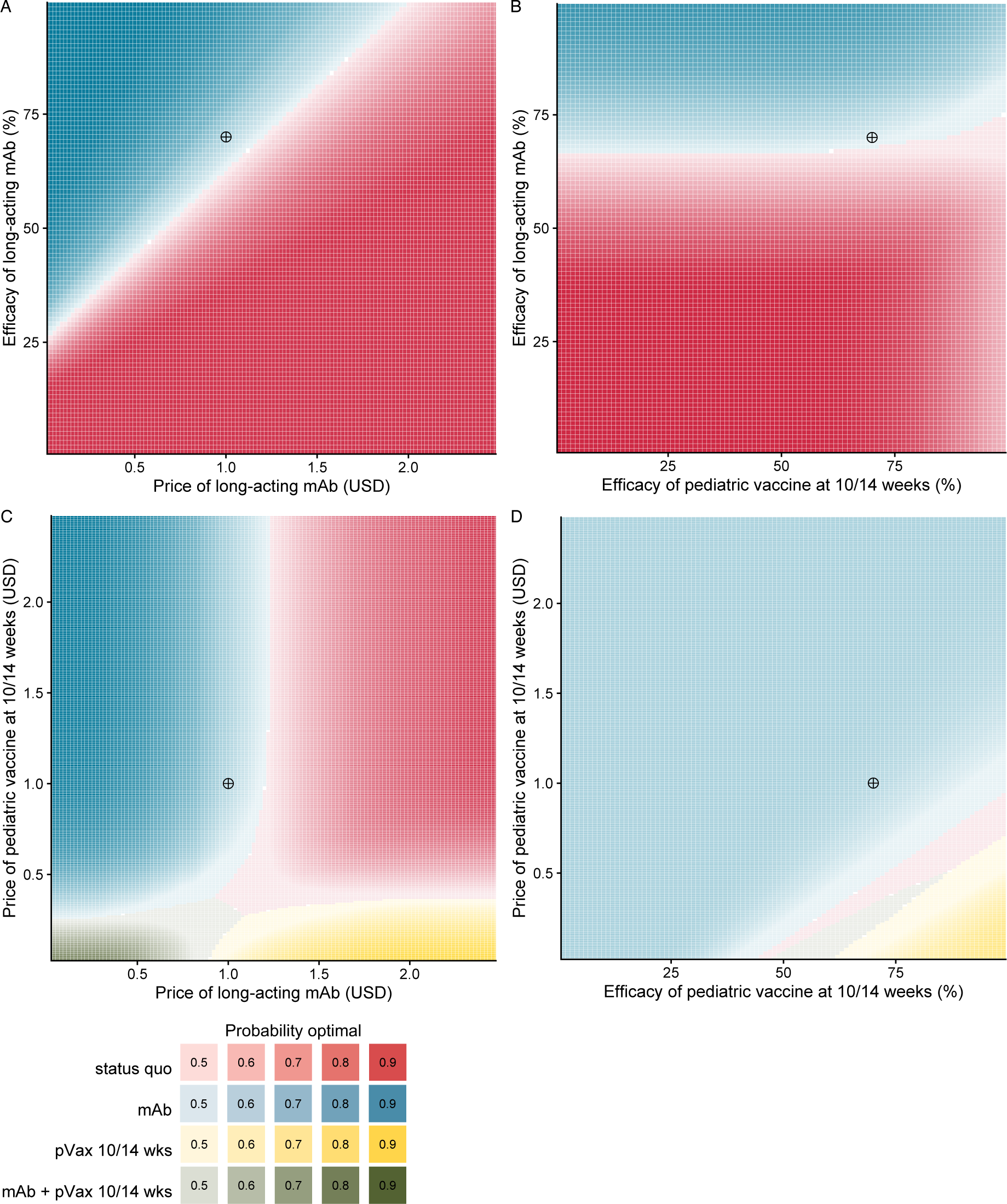
A) Optimal RSV LRTI prevention strategy across changing price of extended half-life mAb and price of pediatric vaccine at 10/14 weeks. B) Optimal RSV LRTI prevention strategy across changing price of extended half-life mAb and efficacy of pediatric vaccine at 10/14 weeks. C) Optimal RSV LRTI prevention strategy across changing price of pediatric vaccine at 10/14 weeks and efficacy of pediatric vaccine at 10/14 weeks. Color saturation is greatest when the optimal strategy choice is most certain. The crosshairs mark the base case across each parameter space.

We conducted an alternative scenario analysis where maternal vaccine has higher coverage and is delivered seasonally as opposed to year-round. These changes did improve the expected value of the maternal vaccine strategy by averting more DALYs, 532 compared to 347 in the base case, and reducing total costs. However, maternal vaccination still did not outperform extended half-life monoclonal antibody and as a result the optimal strategy choice did not change.

## DISCUSSION

We determined that RSV LRTI prevention products would have a substantial impact on early childhood health outcomes in Mali. The seasonal administration of extended half-life mAb immunoprophylaxis at birth would prevent 1300 RSV LRTI hospitalizations, 31 RSV LRTI deaths, and 878 DALYs for children through the first three years of life. In a health economic analysis, we identified that extended half-life mAb prophylaxis is likely to be the optimal next-generation strategy for RSV LRTI prevention in Mali, if the product were priced similarly to routine pediatric vaccines. If extended half-life mAb products were priced below $1.24 per dose, RSV mAb immunoprophylaxis programs in Mali would be cost-effective and optimal compared to the other strategies we evaluated. It may be overly optimistic, however, to assume mAb product would be available at this low price. Gavi currently pays more for other new vaccines, $1.50 per dose for typhoid conjugate vaccine and $2.90 for pneumococcal conjugate vaccine, and the manufacturing costs for mAb are anticipated to be greater than for vaccine products [32]. If mAb was priced above $1.24 per dose, then pediatric RSV vaccines meeting WHO Preferred Product Characteristics would have to be priced below $0.33 to be the optimal strategy option. Above these thresholds, RSV LRTI prevention may not be cost-effective in Mali.

More expensive products may be acceptable if a donor has a WTP that is higher than our base case. Much debate surrounds the appropriate WTP for new interventions in LMICs. Previously used thresholds relied on the per-capita GDP for a country, categorizing interventions as “cost-effective” if they conferred health for an ICER below three times the per capita GDP per DALY averted [34]. These thresholds have come under criticism for neglecting to consider budget impact, affordability, and efficiency [29,30,34].

By contrast, for countries such as Mali with low GDP which rely heavily on donor funding, these thresholds may underestimate donor WTP. For instance, the 2019 per-capita GDP for Mali was $891 [21], which is low even among Gavi-eligible countries, and donors are unlikely to offer fewer product subsidies to the lowest-income countries in their portfolio. A Gavi RSV prevention investment case document from 2018 indicates that the organization’s WTP may be above $1600 [5], indicating a willingness to support prevention in Mali at cost-effectiveness thresholds above those suggested by a Mali-specific analysis. From the government perspective, RSV LRTI prevention becomes optimal at a WTP of $275 per DALY averted, with combination strategies optimal at the GDP-based threshold.

In our study, maternal RSV immunization alone or in combination with other interventions is never the optimal prevention strategy, even in scenarios assuming high maternal vaccine efficacy. The major reason for this is its anticipated four-month duration of protection, shorter than the expected duration for extended half-life mAb [22]. Our model assumed incremental costs of adding maternal immunization to established delivery platforms. In 2020, the Maternal Immunization and Antenatal Care Situation Analysis (MIACSA) study conducted by WHO reported that among 95 LMICs, 44 were categorized as “limited potential to protect mothers and their infants from vaccine preventable diseases” by antenatal care and routine immunization performance [8]. These 44 LMICs were more likely to immunize women through non-routine, supplemental immunization activities [8], indicating the need for substantial system strengthening before routine maternal RSV immunization could be considered. The report implies higher delivery costs than we specified in our study, further eroding the relative efficiency of maternal RSV immunization strategies compared to other interventions. While maternal RSV immunization programs may strengthen antenatal care overall (by supporting health worker salaries or increasing healthcare utilization), these indirect benefits are difficult to quantify and were not incorporated into our analysis.

Similarly, our analysis never identified pediatric vaccination at 6/7 months as the optimal prevention strategy. Although high rates of RSV LRTI persist through the first year of life, the greatest burden of mortality and hospitalization occurs in the first six months. Unfortunately, pediatric RSV vaccines are currently being evaluated for use beginning at six months of age [11]. For use in Mali and similar LMICs, pediatric vaccine products will need to be efficacious when delivered at routine immunization timepoints, typically at 6 to 14 weeks of age.

A major strength of our analysis is that most of our inputs are drawn from studies conducted in Mali. This includes published research on RSV incidence, RSV LRTI hospitalization, and the medical care costs of respiratory infections [1,15–17]. In comparison with studies that evaluate many countries simultaneously, the RSV LRTI incidence we used is somewhat higher and our medical care costs are lower [19,35]. We also specified a base case product price lower than these other analyses, based on products considered acceptable in Mali. With these differences, we find RSV LRTI intervention to be more favorable under a broader range of conditions than similar studies [19].

Our analysis has limitations. First, the RSV incidence inputs came from a single-year, household surveillance study in Bamako. We do not know whether the findings of that study can be generalized to other years or to Mali as a whole. Second, for efficacy and duration, we used WHO Preferred Product Characteristics. While these characteristics were developed by expert committees to be feasible goals for extended half-life mAbs, maternal vaccines, or pediatric vaccines, the absence of licensed products with directly measured performance limits the accuracy of any model outputs. A phase 3 trial of nirsevimab, an extended half-life mAb, showed 74.5% protection against RSV LRTI [36], slightly outperforming the Preferred Product Characteristics. However, this product has an anticipated price point suitable for high-income countries, and effectiveness of the biosimilar products under development may differ [9]. Third, our static model did not account for potential herd immunity effects of RSV LRTI prevention. However, the WHO Preferred Product Characteristics specify short durations of protection, such that indirect effects would be minimal. If durable pediatric vaccines did become available, they would likely be more favorable through the combination of longer direct protection as well as community protection. Fourth, we assumed pediatric vaccine would be delivered in a series of two doses and that there would not be significant protection until after the second dose, similar to influenza vaccines [37]. If a single dose were sufficient, or if there was substantial protection after the first dose, then pediatric vaccination strategies could have greater impact. Finally, our assumption that mAb immunoprophylaxis be delivered seasonally may decrease product and delivery costs, but it would likely cause challenges regarding supply chain, distribution, storage, and health clinic logistics.

Our study provides evidence for the optimal RSV LRTI prevention strategy in Mali, with findings that should be generalizable to many other low-income countries with high RSV LRTI burden. Extended half-life mAbs delivered at birth or integrated into established immunization contacts performed best in our head-to-head comparisons. Maternal RSV vaccination did not fare as well, due to the anticipated limited duration of protection provided by this strategy, and pediatric RSV vaccines at 6/7 months also did not fare as well, due to missing the period of highest hospitalization and mortality. If safe, efficacious, and programmatically suitable RSV LRTI preventive interventions were licensed and affordable to LMICs, our analysis indicates that they would be impactful and cost-effective.

## Data Availability

All data produced are available online at https://github.com/MCFitz/RSV-CEA-combos-Mali.git.

## REFERENCES

1. O’Brien KL, Baggett HC, Brooks WA, et al. Causes of severe pneumonia requiring hospital admission in children without HIV infection from Africa and Asia: the PERCH multi-country case-control study. The Lancet 2019; 394:757–779.

2. Preferred Product Characteristics of Monoclonal Antibodies for Passive Immunization against Respiratory Syncytial Virus (RSV). 2020. Available at: https://www.who.int/immunization/research/ppc-tpp/PPC_RSV-MAbs_Draft_V-0.1-for-consultation.pdf?ua=1. Accessed 30 April 2020.

3. PATH. RSV Vaccine and mAb Snapshot. Available at: https://www.path.org/resources/rsv-vaccine-and-mab-snapshot/. Accessed 4 August 2020.

4. Meeting of the Strategic Advisory Group of Experts on Immunization (SAGE). Available at: http://www.who.int/immunization/sage/meetings/2016/april/en/. Accessed 23 March 2021.

5. Gavi vaccine investment strategy. Available at: https://www.gavi.org/our-alliance/strategy/vaccine-investment-strategy. Accessed 10 May 2020.

6. Williams SR, Driscoll AJ, LeBuhn HM, Chen WH, Neuzil KM, Ortiz JR. National routine adult immunization programs among World Health Organization Member States: An assessment of health systems to deploy future SARS-CoV-2 vaccines. medRxiv. 2020; :2020.10.16.20213967. Available at: https://doi.org/10.1101/2020.10.16.20213967. Accessed 24 March 2021.

7. Recommended Routine Immunizations for Children. Available at: https://www.who.int/immunization/policy/Immunization_routine_table2.pdf. Accessed 24 March 2021.

8. Giles ML, Mantel C, Muñoz FM, et al. Vaccine implementation factors affecting maternal tetanus immunization in low- and middle-income countries: Results of the Maternal Immunization and Antenatal Care Situational Analysis (MIACSA) project. Vaccine 2020; 38:5268–5277. Available at: https://pubmed.ncbi.nlm.nih.gov/32586763/. Accessed 24 March 2021.

9. Ananworanich J, Heaton PM. Bringing Preventive RSV Monoclonal Antibodies to Infants in Low- and Middle-Income Countries: Challenges and Opportunities. Vaccines (Basel) 2021; 9. Available at: https://pubmed.ncbi.nlm.nih.gov/34579198/. Accessed 17 January 2022.

10. Higgins D, Trujillo C, Keech C. Advances in RSV vaccine research and development - A global agenda. Vaccine 2016; 34:2870–2875. Available at: https://pubmed.ncbi.nlm.nih.gov/27105562/. Accessed 23 March 2021.

11. Respiratory Syncytial Virus (RSV) Investigational Vaccine in Infants Aged 6 and 7 Months Likely to be Unexposed to RSV - Full Text View - ClinicalTrials.gov. Available at: https://clinicaltrials.gov/ct2/show/NCT03636906. Accessed 7 April 2022.

12. Glezen WP. Effect of maternal antibodies on the infant immune response. In: Vaccine. Elsevier BV, 2003: 3389–3392. Available at: https://pubmed.ncbi.nlm.nih.gov/12850346/. Accessed 23 March 2021.

13. Shi T, McAllister DA, O’Brien KL, et al. Global, regional, and national disease burden estimates of acute lower respiratory infections due to respiratory syncytial virus in young children in 2015: a systematic review and modelling study. The Lancet 2017; 390:946–958.

14. Laufer RS, Driscoll AJ, Baral R, et al. (In Press) Cost-effectiveness of infant respiratory syncytial virus preventive interventions in Mali: a modelling study to inform policy and investment decisions. 2021;

15. Tapia MD, Sow SO, Tamboura B, et al. Maternal immunisation with trivalent inactivated influenza vaccine for prevention of influenza in infants in Mali: a prospective, active-controlled, observer-blind, randomised phase 4 trial. The Lancet Infectious Diseases 2016; 16:1026–1035.

16. Buchwald AG, Tamboura B, Tennant SM, et al. Epidemiology, Risk Factors, and Outcomes of Respiratory Syncytial Virus Infections in Newborns in Bamako, Mali. Clin Infect Dis 2020; 70:59–66. Available at: http://dx.doi.org/10.1093/cid/ciz157.

17. Orenstein EW, Orenstein LA, Diarra K, et al. Cost-effectiveness of maternal influenza immunization in Bamako, Mali: A decision analysis. PLoS One 2017; 12:e0171499. Available at: http://dx.doi.org/10.1371/journal.pone.0171499.

18. Gavi the Vaccine Alliance. Eligibility. 2020. Available at: https://www.gavi.org/types-support/sustainability/eligibility. Accessed 31 March 2022.

19. Li X, Willem L, Antillon M, Bilcke J, Jit M, Beutels P. Health and economic burden of respiratory syncytial virus (RSV) disease and the cost-effectiveness of potential interventions against RSV among children under 5 years in 72 Gavi-eligible countries. BMC Medicine 2020; 18.

20. Global Burden of Disease Study 2017 (GBD 2017) Disability Weights | GHDx. Available at: http://ghdx.healthdata.org/record/ihme-data/gbd-2017-disability-weights. Accessed 30 April 2020.

21. Mali Data. 2020. Available at: https://data.worldbank.org/country/mali. Accessed 23 April 2020.

22. WHO Preferred Product Characteristics for Respiratory Syncytial Virus (RSV) Vaccines. 2017. Available at: https://www.who.int/immunization/documents/research/who_ivb_17.11/en/. Accessed 4 August 2020.

23. Pathbreaking-Presidential-Healthcare-Reform-Mali.pdf - Google Drive. Available at: https://drive.google.com/file/d/1lH8-MFs7A5qUZ6oMuKhBmscdDib-aXTD/view. Accessed 17 January 2022.

24. PAHO Revolving Fund Vaccine Prices for 2021 - PAHO/WHO | Pan American Health Organization. Available at: https://www.paho.org/en/documents/paho-revolving-fund-vaccine-prices 2021. Accessed 17 January 2022.

25. Baral R, Levin A, Odero C, et al. Costs of continuing RTS,S/ASO1E malaria vaccination in the three malaria vaccine pilot implementation countries. PLOS ONE 2021; 16:e0244995. Available at:https://dx.plos.org/10.1371/journal.pone.0244995. Accessed 17 February 2021.

26. Gavi Board approves funding to support malaria vaccine roll-out in sub-Saharan Africa | Gavi, the Vaccine Alliance. Available at: https://www.gavi.org/news/media-room/gavi-board-approves-funding-support-malaria-vaccine-roll-out-sub-saharan-africa. Accessed 17 January 2022.

27. Fitzpatrick MC, Hampson K, Cleaveland S, et al. Cost-effectiveness of canine vaccination to prevent human rabies in rural Tanzania. Annals of Internal Medicine 2014; 160:91–100. Available at: https://pubmed.ncbi.nlm.nih.gov/24592494/. Accessed 29 March 2021.

28. Stinnett AA, Mullahy J. Net health benefits: a new framework for the analysis of uncertainty in cost-effectiveness analysis. Med Decis Making 1998; 18:S68–80. Available at: http://dx.doi.org/10.1177/0272989x98018002s09.

29. Newall AT, Jit M, Hutubessy R. Are current cost-effectiveness thresholds for low- and middle-income countries useful? Examples from the world of vaccines. Pharmacoeconomics. 2014; 32:525–531. Available at: https://pubmed.ncbi.nlm.nih.gov/24791735/. Accessed 3 November 2020.

30. Jit M. Informing Global Cost-Effectiveness Thresholds Using Country Investment Decisions: Human Papillomavirus Vaccine Introductions in 2006-2018. Value in Health 2021; 24:61–66. Available at:/pmc/articles/PMC7813214/. Accessed 29 March 2021.

31. Antenatal care - UNICEF DATA. Available at: https://data.unicef.org/topic/maternal-health/antenatal-care/. Accessed 4 August 2020.

32. Expanding access to monoclonal antibody-based products: A global c. Available at: https://www.iavi.org/news-resources/expanding-access-to-monoclonal-antibody-based-products-a-global-call-to-action. Accessed 12 April 2022.

33. Tan-Torres Edejer T, Baltussen R, Adam T, et al., editors. Making Choices in Health: WHO Guide to Cost-Effectiveness Analysis. Geneva: World Health Organization: 2003.

34. Bilinski A, Neumann P, Cohen J, Thorat T, McDaniel K, Salomon JA. When cost-effective interventions are unaffordable: Integrating cost-effectiveness and budget impact in priority setting for global health programs. PLOS Medicine 2017; 14:e1002397. Available at: https://dx.plos.org/10.1371/journal.pmed.1002397. Accessed 6 August 2020.

35. Baral R, Higgins D, Regan K, Pecenka C. Impact and cost-effectiveness of potential interventions against infant respiratory syncytial virus (RSV) in 131 low-income and middle-income countries using a static cohort model. BMJ Open 2021; 11.

36. Hammitt LL, Dagan R, Yuan Y, et al. Nirsevimab for Prevention of RSV in Healthy Late-Preterm and Term Infants. New England Journal of Medicine 2022; 386:837–846.

37. Wright PF, Cherry JD, Foy HM, et al. Antigenicity and reactogenicity of influenza A/USSR/77 virus vaccine in children--a multicentered evaluation of dosage and safety. Rev Infect Dis 1983; 5:758–764. Available at: https://pubmed.ncbi.nlm.nih.gov/6353530/. Accessed 29 June 2021.

38. WHO and UNICEF estimates of national immunization coverage. Available at: https://www.who.int/immunization/monitoring_surveillance/data/mli.pdf?ua=1. Accessed 9 May 2020.

39. Frequently Asked Questions (FAQs) on Typhoid Conjugate Vaccine (TCV) support. Available at: https://www.gavi.org/sites/default/files/document/support/Typhoid%20vaccine_FAQ.pdf. Accessed 1 February 2022.

