## Supplementary material for "Optimizing next-generation RSV prevention in Mali: a cost-effectiveness analysis of pediatric vaccination, maternal vaccination, and extended half-life monoclonal antibody immunoprophylaxis": Online Supplement

Contents:

Methods

1. Supplement Table 1: Summary of input parameters used for the base case analysis
2. Supplement Figure 1: Health outcome tree for monthly birth cohorts followed until three years
3. Supplement Figure 2: RSV attack rate probability sampling distributions by age in months
4. Supplement Figure 3: Hospitalization rate given RSV-LRTI probability sampling distributions by age in months
5. Supplement Figure 4: Attributable fatality rate probability sampling distributions by age in months
6. Supplement Figure 5: Comparison of case-fatality rates among hospitalized infants with RSV-LRTI in LMICs and Mali
7. Supplement Figure 6: Other input parameter probability sampling distributions
8. Supplement Figure 7: RSV prevention product administration and protection schedules

Results

1. Supplement Table 2: Budget impact and cost-effectiveness analysis by payer perspective
2. Supplement Figure 8: Optimal strategy across willingness to pay from both the government and donor perspective
3. Supplement Figure 9: Sensitivity analysis on optimal strategy across changing efficacy of the pediatric vaccine at 10/14 weeks

**Supplement Table 1:** Summary of input parameters used for the base case analysis

|  | **Value (95% Confidence Interval, when used)** | **Rationale** |
| --- | --- | --- |
| **Epidemiologic parameters** |  |  |
| RSV attack rate | Age-specific, see Supplement Figure 4. | We started with community-based RSV incidence rates per 1000 person-years in Mali for children in the first six months of life [1]. Incidence varies by infant age as well as calendar month. Incidence rates for Mali were extrapolated out to 36 months based on the linear decline in the RSV incidence rates described for low-income countries between 6 months and 24 months, and between 24 and 36 months [2]. |
| Probability of LRTI given RSV | 0.13 (0.11, 0.17) | All infants with pneumonia episodes occurring between October 2012 and May 2013 were selected to be tested for RSV in the Mali incidence study, whereas only 48.9% of the infants with influenza-like-illness without pneumonia were tested [1]. To account for the oversampling among infants with pneumonia, we calculate the probability of LRTI given RSV as the proportion of RSV cases with pneumonia reported in the trial adjusted to match the proportion of infants with influenza-like-illness but without pneumonia who were tested for RSV. |
| Probability of inpatient care given RSV-LRTI by age | Age-specific, see Supplement Figure 5. | We applied an age-specific gradient for hospitalization rates from the Gambia [3] to the Mali-specific rate of hospitalization given RSV LRTI [1]. Starting at 0·30 in the first month of life, these rates decline to 0·12 by the 36^th^ month of life. All infants with pneumonia who did not receive inpatient care received outpatient care in the Mali community incidence study [1]. The probability of outpatient care given RSV-LRTI is therefore calculated as 1- probability of inpatient care given RSV-LRTI. |
| Case fatality rate among those who received inpatient care given RSV-LRTI by age | Age-specific, see Supplement Figure 6. | We used age-specific hospital case fatality rates for RSV LRTI in LMICs [4]. The probability of death given inpatient care is 0·27 in the first month of life and declines to 0·008 by the 36^th^ month of life. We assume 49% of RSV-LRTI deaths occur outside the hospital [3]. |
| Disability weight inpatient RSV-LRTI | 0.13 (0.10, 0.17) | The 2017 Institute for Health Metrics and Evaluation Global Burden of Disease disability weight for an acute episode of a severe LRTI, used to calculate DALYs [5]. |
| Disability weight outpatient RSV-LRTI | 0.05 (0.04, 0.07) | The 2017 Institute for Health Metrics and Evaluation Global Burden of Disease disability weight for an acute episode of a moderate LRTI, used to calculate DALYs [5] |
| Duration of RSV illness (days) | 8.5 (7, 10) | The average duration of illness for episodes of lower-respiratory tract infections based on health facility data [6]. |
| **Demographic parameters** |  |  |
| Crude birth rate (per 1,000) | 42 | The World Bank crude birth rate per 1,000 total population for Mali in 2017 [7]. |
| Total country population | 18,540,000 | The World Bank total population estimate for Mali in 2017 [7]. |
| Number of infants in each birth cohort | 59,734 – 66,134 | The number of infants in each monthly birth cohort was calculated by first multiplying the crude birth rate by the total country population for Mali to estimate the number of infants born in one year. The total number of births for each month was assigned based on the number of days in each month. |
| Life expectancy at birth | 58 | Average 2017 life expectancy at birth for Mali, used to calculate DALYs [7]. |
| **Economic parameters** |  |  |
| Inpatient care costs (USD) | 118.57 (92.20, 144.68) | Average cost of medical care for infants less than six months with confirmed RSV illness who received inpatient care. Costs are inclusive of outpatient services also acquired by this group [8]. We assume 53% of severe RSV-LRTI episodes do not receive inpatient care [3]. |
| Outpatient care costs (USD) | 6.56 (5.44, 7.66) | Average cost of medical care for infants less than six months with confirmed RSV illness who received outpatient care services only [8]. |
| Administration cost of adding a new product to a childhood immunization visit (USD) | 0.67 | The mean incremental cost of adding one product to the established immunization schedule in low-income countries, equal to $0.67 [9]. |

**Supplement Figure 1:** Health outcome tree for monthly birth cohorts followed until three years

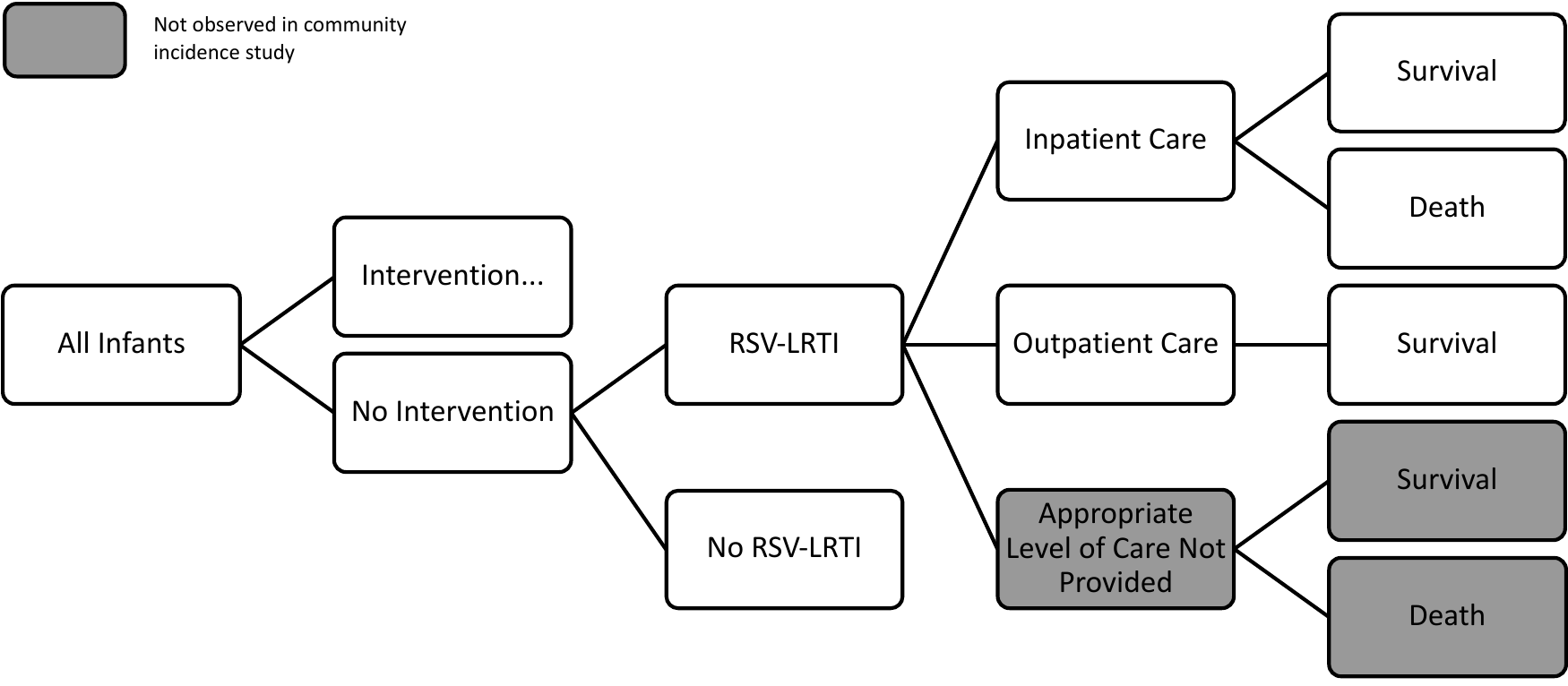

**Supplement Figure 2:** RSV attack rate probability sampling distributions by month of age. The red line represents the point estimate.

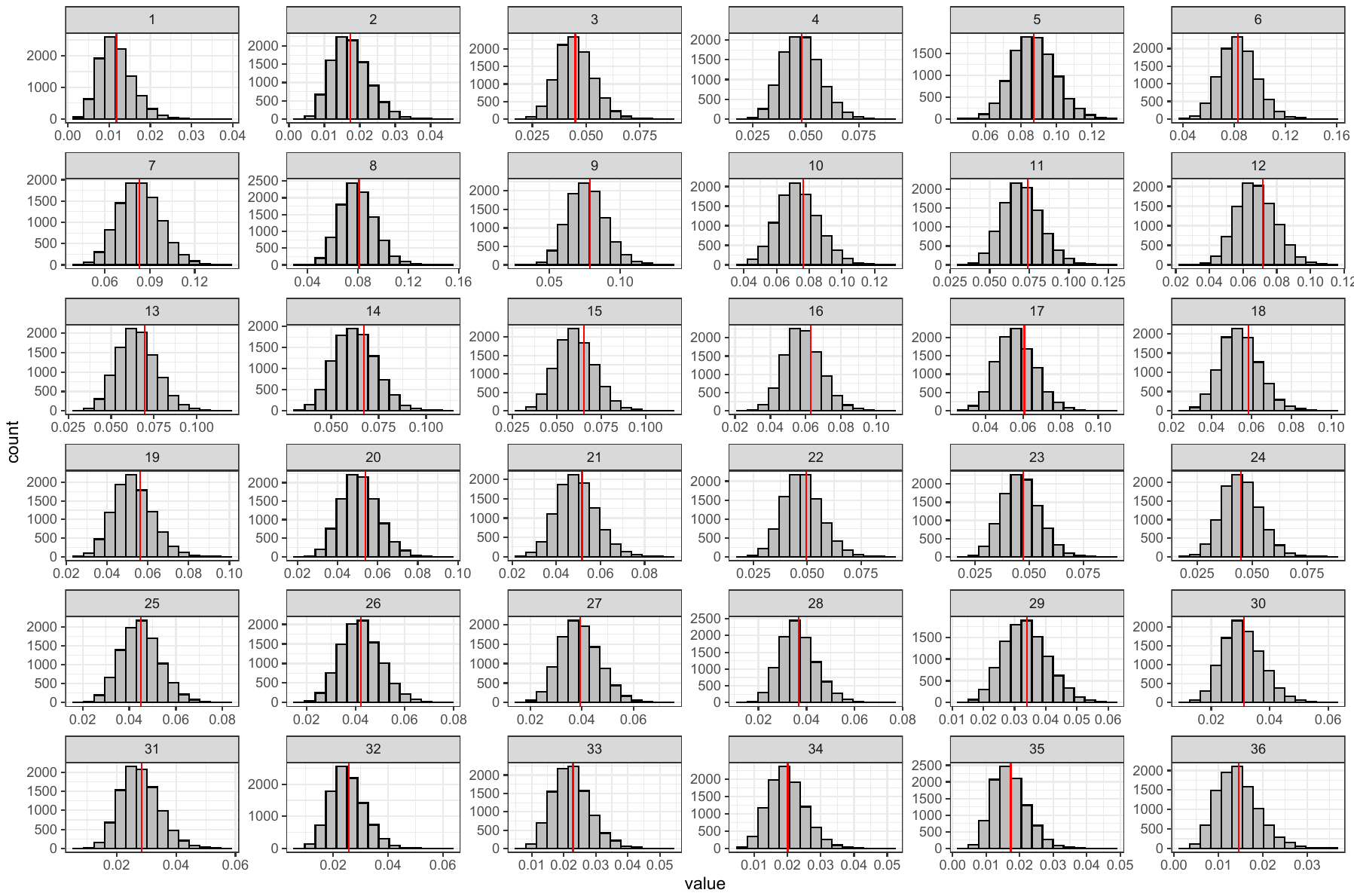

**Supplement Figure 3:** Hospitalization rate given RSV-LRTI probability sampling distributions by age. The red line represents the point estimate.

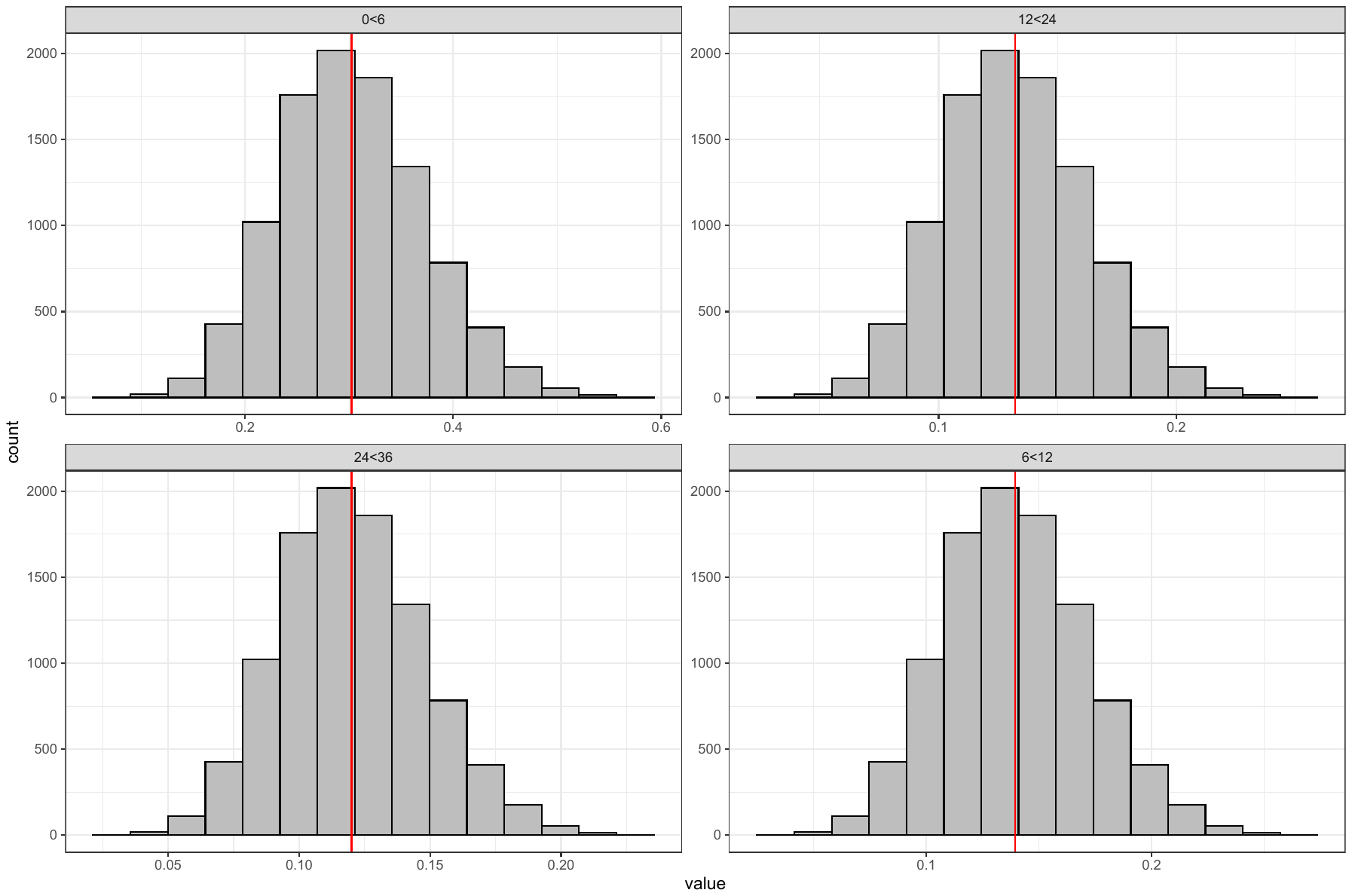

**Supplement Figure 4:** Attributable fatality rate given RSV-LRTI hospitalization probability sampling distributions by month of age. The red line represents the point estimate.

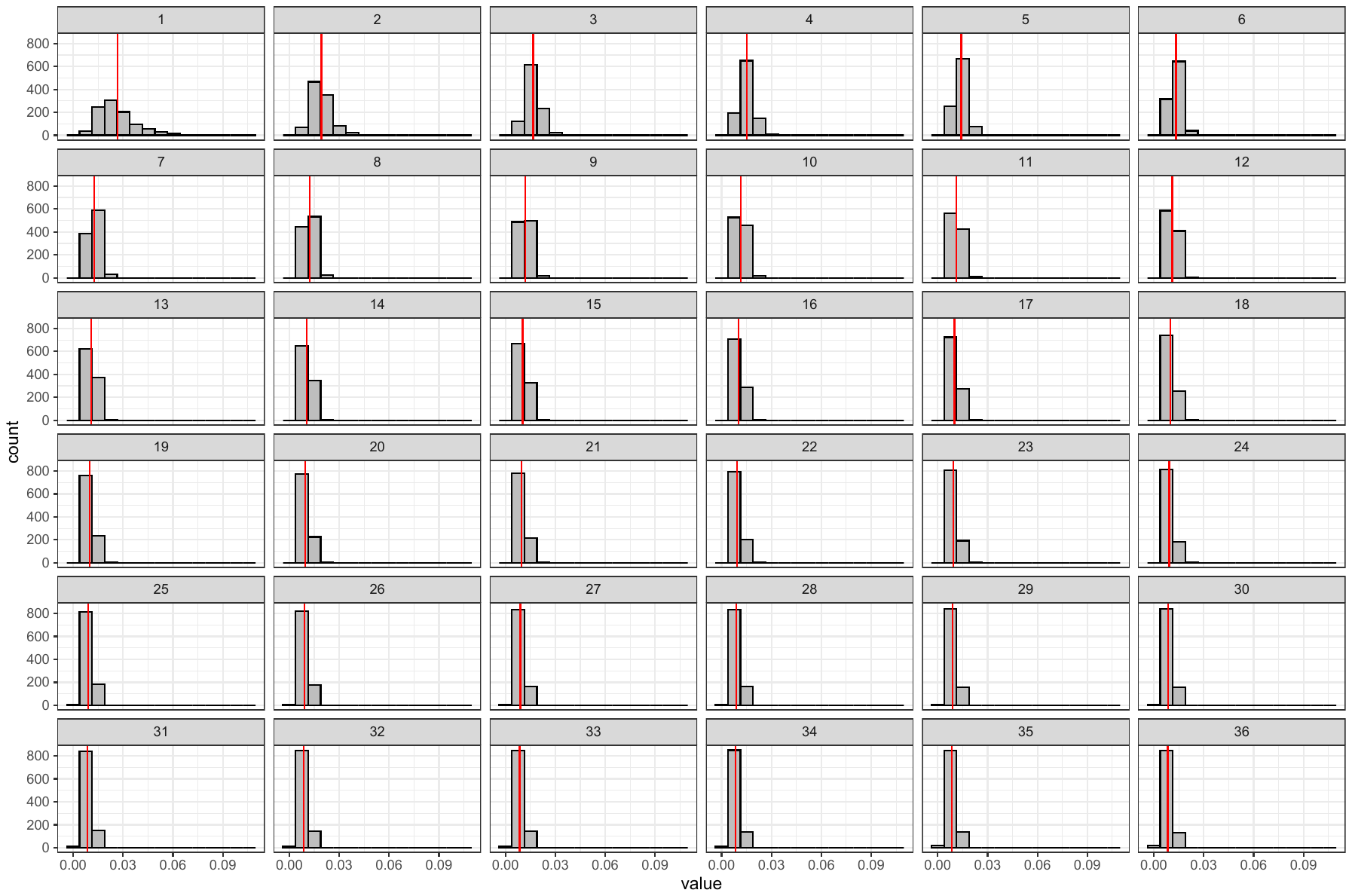

**Supplement Figure 5:** Comparison of case-fatality rates (CFR) in the first six months of life among hospitalized infants with RSV-LRTI in LMICs [3], and Mali [10]. T-bars indicate the 95% confidence intervals.

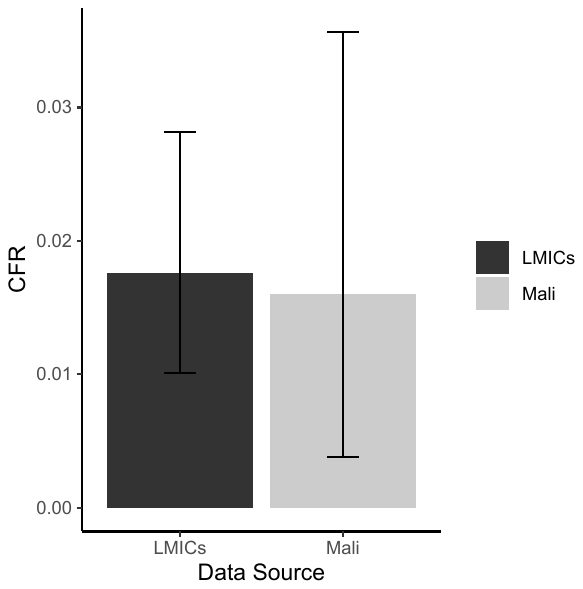

**Supplement Figure 6:** Other input parameter probability sampling distributions. The red line represents the point estimate.

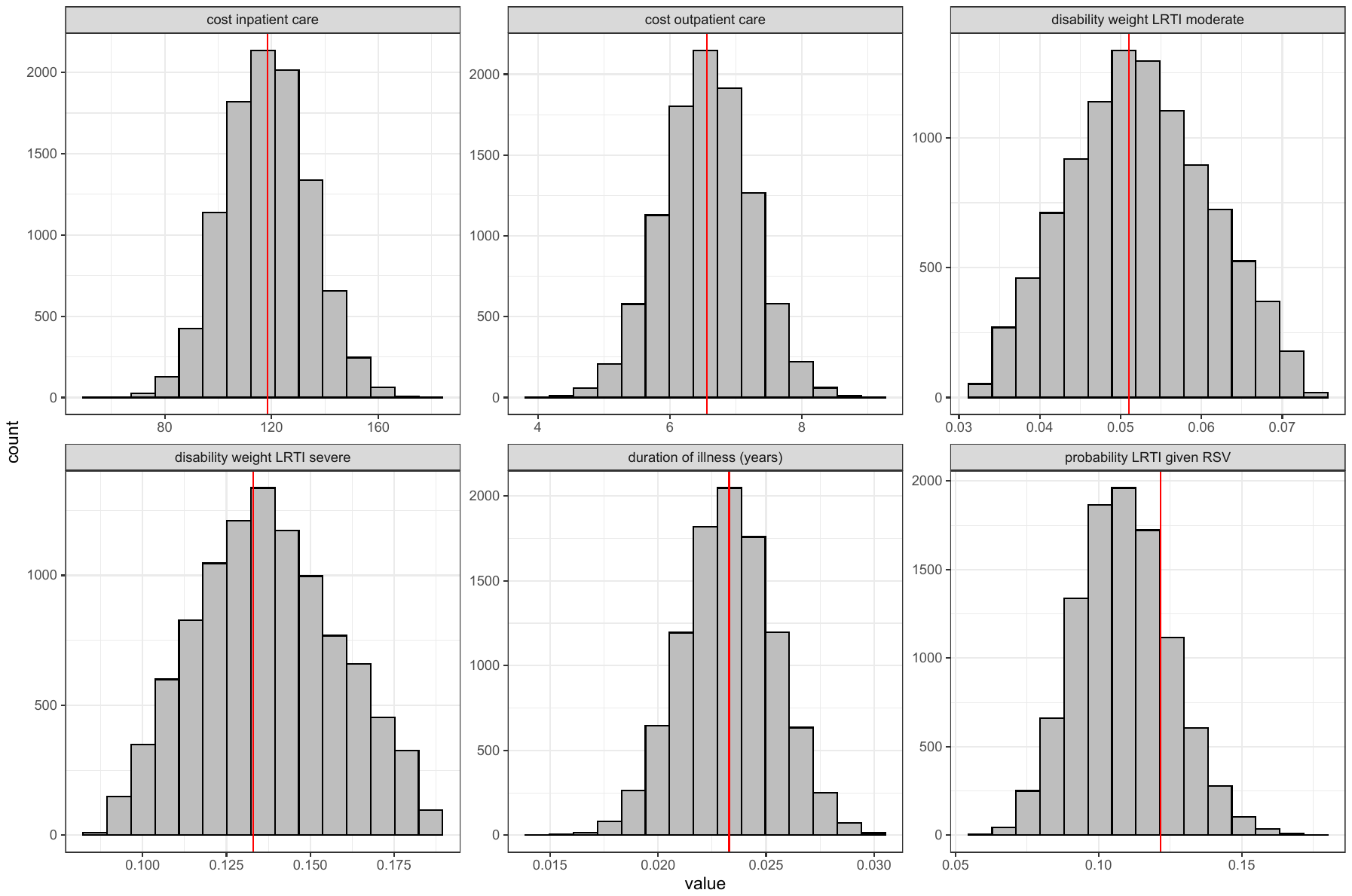

**Supplement Figure 7:** RSV prevention product administration and protection schedules

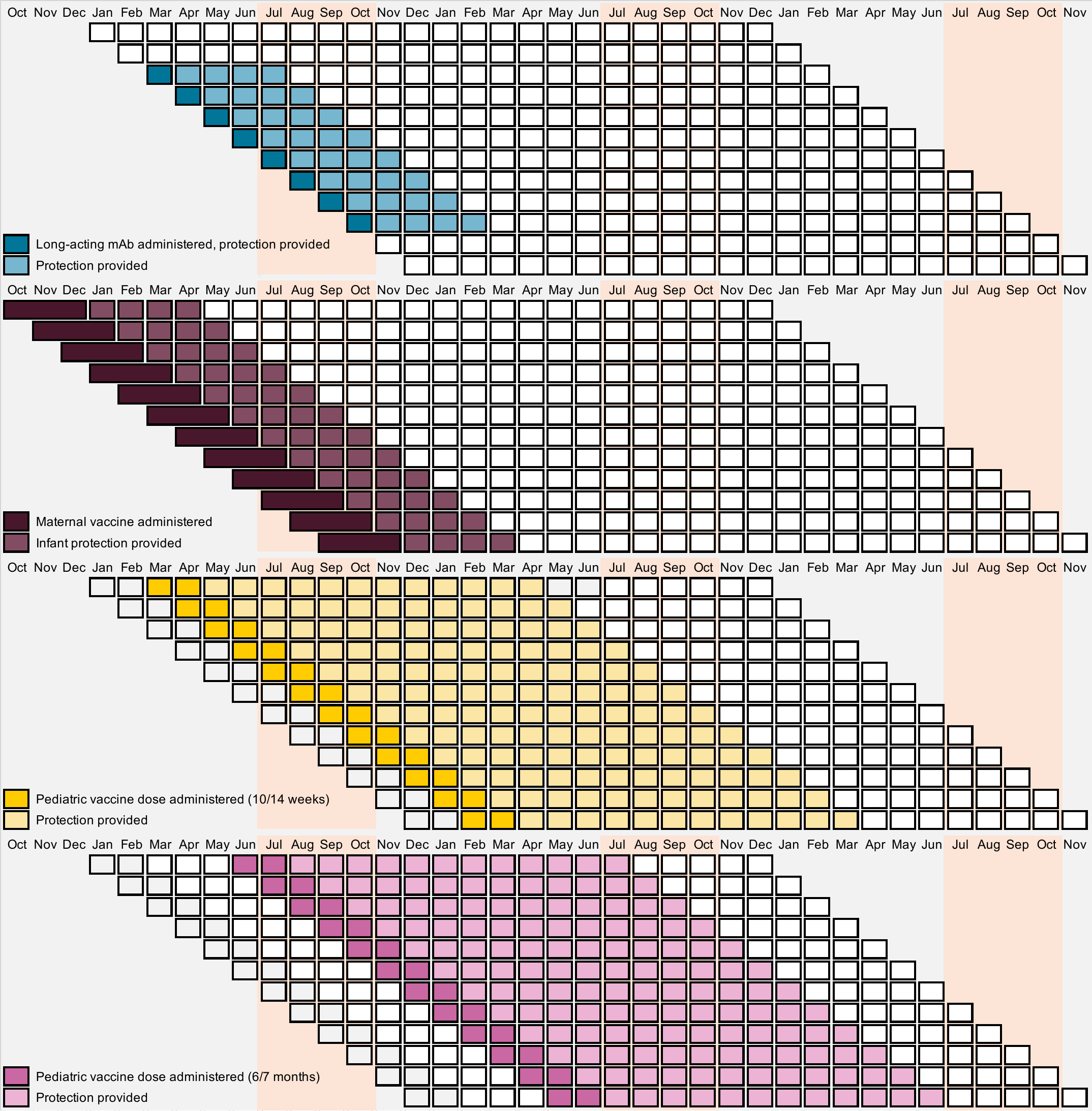

**Supplement Table 2:** Budget impact and cost-effectiveness analysis by payer perspective

|  | DALYs averted compared to status quo | Medical costs | Donor costs | Incremental cost-effectiveness ratio, donor perspective | Government costs | Incremental cost-effectiveness ratio, government perspective | Societal costs | Incremental cost-effectiveness ratio, societal perspective |
| --- | --- | --- | --- | --- | --- | --- | --- | --- |
| Status quo | --- | $1249382 (742201, 1546908) | --- | --- | $1249382 (742201, 1546908) | --- | $1249382 (742201, 1546908) | --- |
| Maternal vaccine | 347 (140, 569) | $1176679 (699904, 1455074) | $269954 | *Weakly dominated* | $1470194 (977962, 1719329) | *Weakly dominated* | $1740148 (1263373, 2018543) | *Weakly dominated* |
| Long-acting monoclonal antibody | 878 (369, 1393) | $1048241 (624702, 1288465) | $347339 | $396 (50, 928) | $1425895 (989196, 1652832) | $201 (94, 664) | $1773233 (1349695, 2013457) | $597 (354, 1621) |
| Pediatric vaccine at 10 & 14 weeks | 1247 (548, 1848) | $817669 (486934, 1018490) | $960114 | *Weakly dominated* | $1861579 (1523004, 2045810) | *Weakly dominated* | $2821693 (2490958, 3022514) | *Weakly dominated* |
| Pediatric vaccine at 6 & 7 months | 558 (245, 805) | $1006701 (596633, 1261419) | $872831 | *Strongly dominated* | $1955710 (1532775, 2198145) | *Strongly dominated* | $2828541 (2418472, 3083259) | *Strongly dominated* |
| Maternal vaccine plus sequential pediatric vaccine at 10 & 14 weeks | 1594 (705, 2387) | $744966 (445934, 924936) | $1230068 | *Weakly dominated* | $2082391 (1774726, 2245343) | *Weakly dominated* | $3312459 (3013427, 3492429) | *Weakly dominated* |
| Maternal vaccine plus sequential pediatric vaccine at 6 & 7 months | 906 (400, 1343) | $933998 (554562, 1167781) | $1142785 | *Strongly dominated* | $2176522 (1784383, 2397700) | *Strongly dominated* | $3319307 (2939871, 3553090) | *Strongly dominated* |
| Long-acting monoclonal antibody plus sequential pediatric vaccine at 6 & 7 months | 1436 (630, 2155) | $805560 (480774, 1002645) | $1220169 | *Strongly dominated* | $2132223 (1795509, 2319766) | *Strongly dominated* | $3352392 (3027606, 3549477) | *Strongly dominated* |
| Long-acting monoclonal antibody plus sequential pediatric vaccine at 10 & 14 weeks | 1947 (855, 2936) | $662438 (398664, 819499) | $1307452 | $898 (605, 2034) | $2084002 (1812017, 2225676) | $615 (380, 1660) | $3391454 (3127680, 3548516) | $1514 (999, 3705) |

**Supplement Figure 8:** **A)** Optimal strategy across willingness to pay from the government perspective. **B)** Optimal strategy across willingness to pay from the donor perspective.

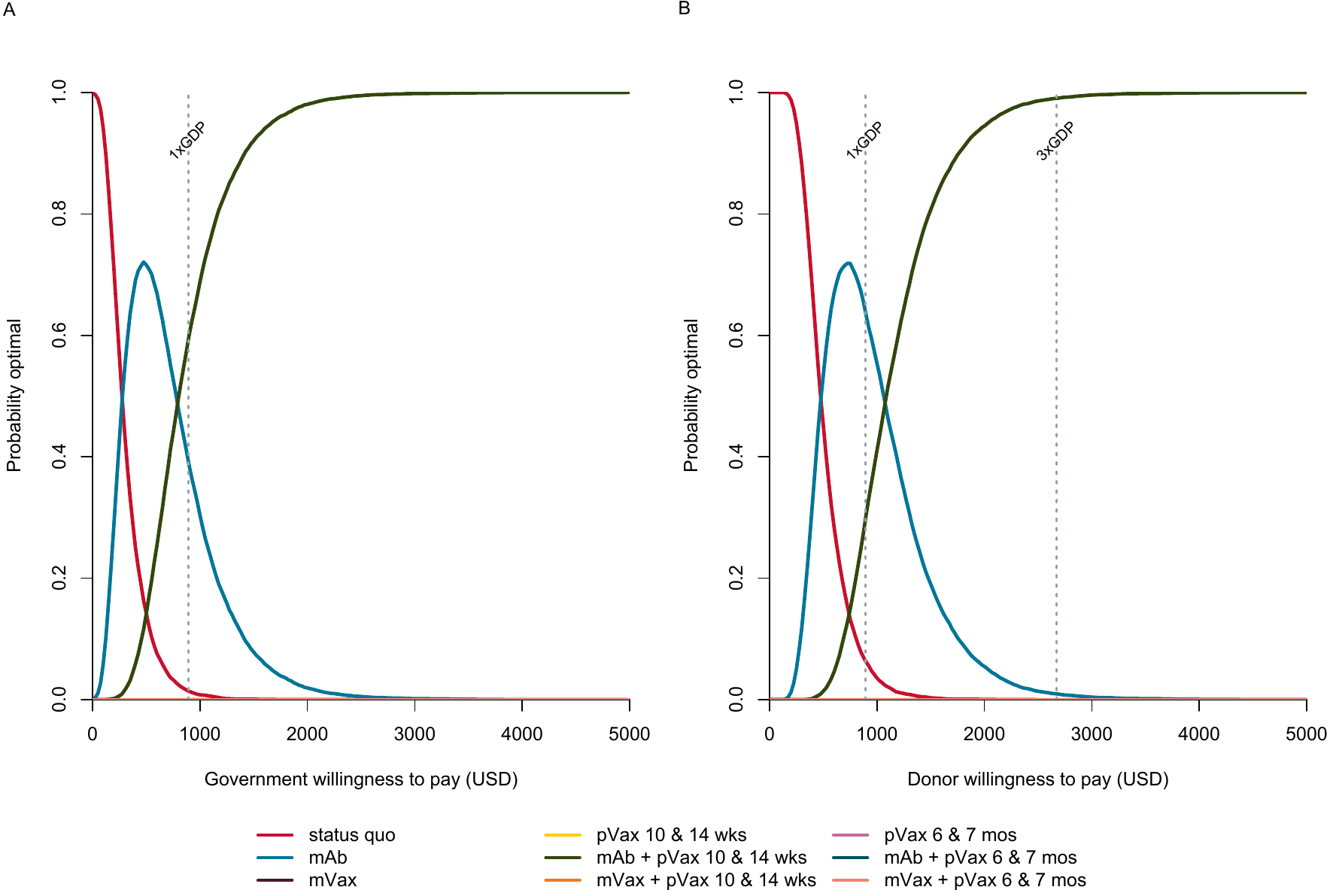

**Supplement Figure 9: A)** Sensitivity analysis on optimal strategy across changing efficacy of the pediatric vaccine when administered at 10/14 weeks. **B)** Sensitivity analysis on optimal strategy across changing efficacy of the pediatric vaccine when administered at 10/14 weeks as part of a combination strategy in which either maternal vaccine or monoclonal antibodies were administered prior to the pediatric vaccine.

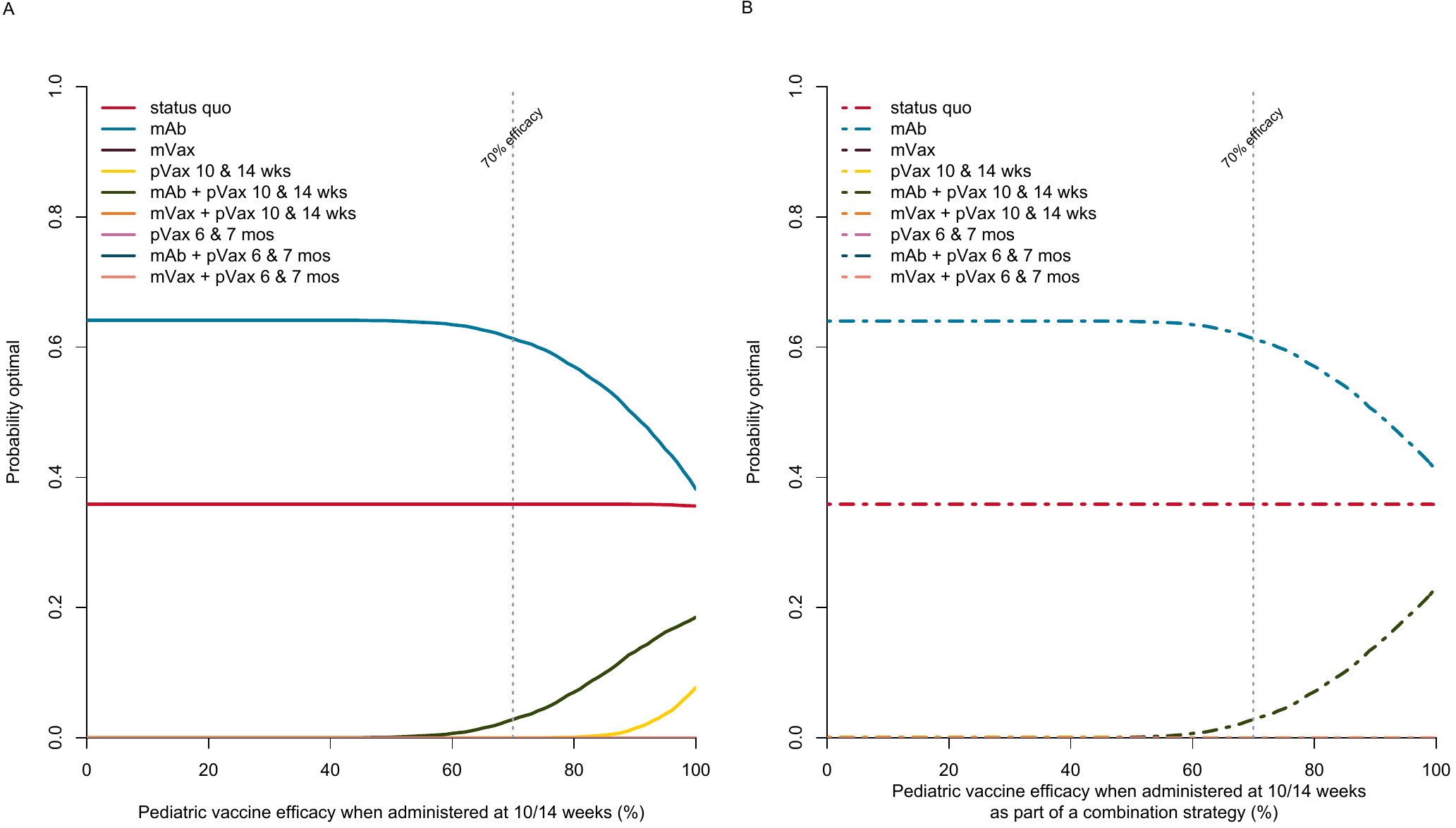

**References:**

1.	Buchwald AG, Tamboura B, Tennant SM, et al. Epidemiology, Risk Factors, and Outcomes of Respiratory Syncytial Virus Infections in Newborns in Bamako, Mali. Clin Infect Dis **2020**; 70:59–66. Available at: http://dx.doi.org/10.1093/cid/ciz157.

2.	Li X, Willem L, Antillon M, Bilcke J, Jit M, Beutels P. Health and economic burden of respiratory syncytial virus (RSV) disease and the cost-effectiveness of potential interventions against RSV among children under 5 years in 72 Gavi-eligible countries. BMC Medicine **2020**; 18.

3.	Shi T, McAllister DA, O’Brien KL, et al. Global, regional, and national disease burden estimates of acute lower respiratory infections due to respiratory syncytial virus in young children in 2015: a systematic review and modelling study. The Lancet **2017**; 390:946–958.

4.	Li X, Willem L, Antillon M, Bilcke J, Jit M, Beutels P. Health and economic burden of respiratory syncytial virus (RSV) disease and the cost-effectiveness of potential interventions against RSV among children under 5 years in 72 Gavi-eligible countries. BMC Med **2020**; 18:82. Available at: https://www.ncbi.nlm.nih.gov/pubmed/32248817.

5.	Global Burden of Disease Study 2017 (GBD 2017) Disability Weights | GHDx. Available at: http://ghdx.healthdata.org/record/ihme-data/gbd-2017-disability-weights. Accessed 30 April 2020.

6.	Mathers CD, Vos T, Lopez AD, Salomon J, Ezzati M. National Burden of Disease Studies: A Health, Practical Guide. Edition 2.0. Global Program on Evidence for Health Policy. Geneva: World Health Organization: 2001. Available at: https://www.who.int/healthinfo/nationalburdenofdiseasemanual.pdf.

7.	Mali Data. 2020. Available at: https://data.worldbank.org/country/mali. Accessed 23 April 2020.

8.	Orenstein EW, Orenstein LA, Diarra K, et al. Cost-effectiveness of maternal influenza immunization in Bamako, Mali: A decision analysis. PLoS One **2017**; 12:e0171499. Available at: http://dx.doi.org/10.1371/journal.pone.0171499.

9.	Baral R, Higgins D, Regan K, Pecenka C. Impact and cost-effectiveness of potential interventions against infant respiratory syncytial virus (RSV) in 131 low-income and middle-income countries using a static cohort model. BMJ Open **2021**; 11:46563. Available at: http://bmjopen.bmj.com/. Accessed 17 January 2022.

10.	O’Brien KL, Baggett HC, Brooks WA, et al. Causes of severe pneumonia requiring hospital admission in children without HIV infection from Africa and Asia: the PERCH multi-country case-control study. The Lancet **2019**; 394:757–779.
